# ABCDEF Bundle Implementation: The influence of access to bundle-enhancing supplies and equipment

**DOI:** 10.1101/2021.05.13.21257046

**Authors:** Alvin D. Jeffery, Jennifer A. Werthman, Valerie Danesh, Mary S. Dietrich, Lorraine C. Mion, Leanne M. Boehm

**Affiliations:** Geriatric Research, Education and Clinical Center (GRECC), VA Tennessee Valley Healthcare System, Nashville, TN, USA; School of Nursing, Vanderbilt University, Nashville, TN, USA; Department of Biomedical Informatics, Vanderbilt University Medical Center, Nashville, TN, USA; Baylor Scott & White Health Research Institute, Dallas, TX, USA; School of Nursing, University of Texas at Austin, Austin, TX; College of Nursing, The Ohio State University, Columbus, OH, USA; Center for Healthy Aging, Self Management, and Complex Care, The Ohio State University, Columbus, OH, USA; Critical Illness, Brain Dysfunction, and Survivorship Center, Vanderbilt University Medical Center, Nashville, TN, USA

## Abstract

**Importance:** The ABCDEF bundle is a guideline-recommended framework for implementing evidence-based practices in the Intensive Care Unit (ICU), but it is underutilized across the world.

**Objective:** Describe the physical environment factors (i.e., availability, accessibility) of bundle-enhancing items in units implementing the bundle and the influence of physical environment on bundle adherence.

**Design, Setting, and Participants:** This multicenter, exploratory, cross-sectional study used data from two ICU-based randomized controlled trials (RCTs) (NCT01211522 and NCT01739933) that measured daily bundle adherence. The study included 10 medical and surgical ICUs in 6 academic medical centers in the continental United States. Adults with qualifying respiratory failure and/or septic shock (e.g., mechanical ventilation, vasopressor use) were included in the RCTs. Unit- and patient-level data collection occurred between 2011 and 2016. We conducted hierarchical logistic regression models using Frequentist and Bayesian frameworks.

**Exposure:** The ABCDE bundle (<u>A</u>wakening and <u>B</u>reathing trial <u>C</u>oordination, <u>D</u>elirium assessment/management, <u>E</u>arly mobility) was recommended standard of care for RCT patients and adherence tracked daily.

**Measure(s):** The primary outcome was adherence to the full bundle and the early mobility bundle component as identified from daily adherence documentation (n=751 patient observations). Unit-level measures included minimum and maximum distances to 25 bundle-enhancing items and the relationship to bundle adherence.

**Results:** In all cases, mechanical ventilation was associated with decreased bundle adherence. Some of the models suggested the following variables were also influential: age (older associated with decreased adherence), unit size (larger associated with decreased adherence), and a standard walker (presence associated with increased adherence).

**Conclusions and Relevance:** Both unit- and patient-level barriers influenced full bundle and early mobility implementation. There is potential benefit of physical proximity to essential items for ABCDEF bundle and early mobility adherence. Future studies with larger sample sizes should explore how equipment location and availability influences practice.

**Key Points:** *Question:* Does the physical environment, specifically the availability and accessibility of ABCDEF bundle-enhancing items, influence bundle adherence?

*Findings:* In this cross-sectional study in 10 units evaluating ABCDEF bundle adherence across 751 patient observations, units with access to a standard walker were significantly more likely to provide bundled care. Across all models, patients who were on the ventilator or older were significantly less likely to received bundled care.

*Meaning:* Both unit- and patient-level factors influence ABCDEF bundle implementation and amenable targets for implementation strategy development.

## Introduction

The ABCDEF bundle (**<u>A</u>**ssess, prevent and manage pain; **<u>B</u>**oth spontaneous awakening and breathing trials; **<u>C</u>**hoice of analgesia/sedation; **<u>D</u>**elirium: assess, prevent, and manage; **<u>E</u>**arly mobility; **<u>F</u>**amily engagement and empowerment) is a guideline-recommended, evidence-based approach to organizing Intensive Care Unit (ICU) people, processes, and technology for improved care of the critically ill patient.^1-3^ ABCDEF bundle implementation is associated with reduced delirium, ventilator, ICU, and hospital days; improved survival; and increased likelihood of early mobilization and restraint-free care.^4-6^ Despite guideline recommendations, the bundle is still underutilized in ICUs across the world.^7^ Organizational structure and process factors (e.g., physical environment, staffing, autonomy, workload) have commonly been identified as barriers to bundle implementation.^8-11^

Reason’s Human Errors conceptualization recognizes the influence of latent conditions (i.e., dormant conditions that only become evident with triggering factors).^12^ In the context of ABCDEF implementation, the latent conditions of the physical environment may influence behavioral outcomes. The physical environment, or physical space, of the ICU setting includes the nursing unit configuration, building-related characteristics (e.g., fixtures, dialysis connection placements, distance to equipment storage rooms), and the built environment of modifiable characteristics such as the presence of ceiling lifts, the reach distance from bed to tray table, and the visibility of mobility equipment.

The relationship between the physical environment and care delivery variability has been explored in diverse healthcare settings and patient populations. For example, studies of furniture placement proximity affect interpersonal distance and subsequent self-disclosure during patient-provider history interviews.^13^ In acute care settings, time-motion studies assessing the variability of care between hospitalized patients with and without contact isolation precautions suggest that donning additional isolation gowns with each patient room entry are associated with adverse effects.^14^ Adverse effects of contact isolation include care variability such as decreases in patient-clinician contact and decreases in patient satisfaction.^15, 16^ Variability associated with the change in the physical environment (isolation gowning requirement) is associated with increases in non-infectious adverse events and higher rates of medication errors.^15, 17, 18^ To date, associations between the physical environment and ABCDEF implementation uptake at the nursing shift-level has not been explored.

The aim of this study is to describe the physical environment factors (i.e., availability, accessibility) of ABCDEF bundle-enhancing items in units implementing the bundle and the influence of physical environment on bundle adherence.

## Materials and Methods

This is an exploratory multicenter cross-sectional study conducted with sites participating in two ICU-based randomized controlled trials (RCTs) (NCT01211522 and NCT01739933). Data collection occurred between 2011 and 2016. At the initiation of both studies, the bundle was still an evolving framework for critical care open to evidence-based modification. At the time of these RCTs, we applied the original ABCDE bundle framework (<u>A</u>wakening and <u>B</u>reathing trial <u>C</u>oordination, <u>D</u>elirium assessment/management, <u>E</u>arly mobility) and use this term for our materials and methods description.^19, 20^ The bundle has since been modified to include current critical care guidelines (i.e., ABCDEF bundle, see www.iculiberation.org and www.icudelirium.org).^1, 2^ Ethical approval was obtained from the institutional review board at each of the participating centers.

### Setting & Sample

We obtained information from medical and surgical ICUs (n=10) at six academic medical centers across the continental United States. Hospital size ranged from 175 to 1,541 licensed beds and 10 to 40 beds per ICU. Size of participating units ranged from 987 to 3,412 square meters (m^2^) (median=1,981 m^2^).

Prospective observational patient data was obtained from two ICU-based randomized controlled trials (RCTs) (NCT01211522 and NCT01739933) that employed the bundle as standard of care and measured daily bundle adherence.^21, 22^ Adults with qualifying respiratory failure and/or septic shock (e.g., mechanical ventilation, vasopressor use) were included in the RCTs. Patients with severe cognitive impairment, drug allergy (e.g., haloperidol, propofol), moribund state, cardiac arrythmias (i.e., Torsades de Pointes, 2^nd^ or 3^rd^ degree heart block), unable to speak or understand English, or incarcerated were excluded. Patient screening, randomization, follow-up and analysis are published in the parent RCT reports.^21, 22^

### Variables and Measures

We generated a comprehensive list of supplies and equipment (25 items) for the completion of ABCDE bundle components through communication with clinicians providing critical care. We measured the minimum and maximum distances (in meters) from a head of bed closest to and farthest from each of the 25 bundle-enhancing items.

ABCDE bundle adherence is defined as completing all 5 components on ICU days requiring mechanical ventilation (MV) and completion of delirium assessment/management and early mobility (DE) components on MV-free days. We calculated full bundle adherence for the entire period of MV ICU days as [(days of full bundle adherence)/(total MV ICU days)].^9^ We independently calculated adherence to the E bundle component on MV days and MV-free days using the same equation.

### Procedures

The principal investigator (LMB) personally visited each site to meet individually with the leadership of participating units and collect measurements. LMB operated a measuring wheel to capture exact distances for the minimum and maximum distances from the closest and farthest head of bed to each of the 25 bundle-enhancing items.

The ICU teams completed the ABCDE bundle at their discretion, guided by a standardized protocol as part of the parent RCTs.^21, 22^ The investigators were not responsible for ABCDE bundle performance. An ABCDE bundle checklist was placed at the patient’s bedside and completed by the nurse or other healthcare professional each calendar day for RCT participants.^9^ Study staff distributed, collected, and recorded checklists daily. We collected and managed all study data using Research Electronic Data Capture (REDCap) tools hosted at Vanderbilt University.^23^

### Statistical Methods

We developed two hierarchical logistic regression models using Frequentist and Bayesian frameworks with predictors comprising whether a patient was on a ventilator on a given day, the patient’s age and body mass index, and geospatial measurements (unit size and distances to 25 unique pieces of equipment). The first assumed a binary present/absent method for unit equipment. The second included the actual distances from head of bed to specific unit equipment. In all models, we nested patient-days within patients and patients within units (i.e., 3-level hierarchy).

We modeled two primary outcomes: (1) adherence with the full bundle and (2) adherence with the early mobility [E] component, given it had the largest variance of all components and 23 of the 25 items were directly associated with early mobility. For units that did not have a piece of equipment, we imputed the missing values with two methods: (1) representing missing values as 0 and observed values as 1 – the *binary* models and (2) replacing missing values with twice the maximum observed value to conceptually represent the equipment being even farther away – the *continuous* models. We imputed missing data for height (5.7% missing) and weight (4.8% missing) using the IterativeImputer function from Python’s scikit-learn library. As a final pre-processing step, we scaled all non-binary, numeric variables to have a mean of 0 and variance of 1. Given the large number of predictors (p=29) for the relatively small number of patient observations (n=751), we conducted a redundancy analysis using subject matter expertise, examining bivariate correlations coefficients, and building regression models to determine which variables could be predicted from remaining variables. This dimensionality reduction process resulted in the inclusion of 6 predictors in the binary models and 7 predictors in the continuous models (see eFigures 1-4 and eTable 1 in the Supplement for further details).

**Table 1:** Descriptive statistics patient- and unit-level variables

| **Patient Variables**<br>**(N=105)** | **N**<br>**(% missing)** | **Mean (SD)** | **Median (IQR)** | **Included in**<br>**Final**<br>**Analysis** |
| --- | --- | --- | --- | --- |
| Age | 0 (0) | 54 (14) | 57 (47,63) | Yes |
| Height (cm) | 6 (6) | 171 (12) | 173 (164,178) | Yes |
| Weight (kg) | 5 (5) | 100 (38) | 95 (75,115) | Yes |
| BMI | 6 (6) | 35 (17) | 32 (25,38) | Yes |
| **Unit Variable (N=10)** | **N**<br>**(% missing)** | **Mean (SD)** | **Median (IQR)** | **Included in**<br>**Final**<br>**Analysis** |
| Unit Size (m<sup>2</sup>) | 10 (0) | 2,088 (1,981) | 1,981 (1,103;<br>3,109) | Yes |
| **Distance from HOB to**<br>**Bundle-**<br>**Enhancing Items** | **N units with**<br>**item**<br>**(% missing)** | **Mean (SD)**<br>**(meters)** | **Median (IQR)**<br>**(meters)** | **Included in**<br>**Final**<br>**Analysis** |
| Bag valve mask | 10 (0) | 1.5 (0.7) | 1.4 (1.1, 1.6) |  |
| Bariatric chair | 2 (80) | 67.2 (4.4) | 67.2 (65.6, 68.7) | Yes |
| Canvas sling | 8 (20) | 52.8 (9.5) | 51.2 (46.5, 59) | Yes |
| Ceiling lift | 7 (30) | 1.3 (0.3) | 1.3 (1, 1.7) | Yes |
| Chart | 10 (0) | 15.16 (9.8) | 11.7 (10, 14.9) |  |
| Gait belt | 3 (70) | 50.3 (16.7) | 40.9 (40.6, 55.3) |  |
| High back chair | 5 (50) | 50.4 (21.5) | 64.4 (30.7, 66.8) | Yes |
| Hover mat | 4 (60) | 68.2 (54.4) | 68.2 (49, 87.4) |  |
| Lift sheet | 4 (60) | 17.7 (1.1) | 17.7 (17.3, 18.1) |  |
| Medication dispensing<br>system | 10 (0) | 26.4 (5.8) | 26.4 (21.8, 29.6) |  |
| Nonpharmacologic aids<sup>a</sup> | 2 (80) | 40.6 (0.4) | 40.6 (40.5, 40.8) | Yes |
| Oxygen tank | 10 (0) | 42 (13.2) | 42.4 (30.3, 49.8) | Yes |
| Oxygen tubing | 10 (0) | 50.2 (14.6) | 50.5 (43.5, 57.5) | Yes |
| PEEP valve | 10 (0) | 50.2 (14.6) | 50.5 (43.5, 57.5) | Yes |
| Portable monitor | 2 (80) | 50.8 (18.7) | 50.8 (44.2, 57.5) | Yes |
| Portable ventilator | 2 (80) | 46.1 (2) | 46.1 (45.4, 46.8) | Yes |
| Radio | 1 (90) | 0.9 (NA) | 0.9 (0.9, 0.9) |  |
| Recliner chair | 7 (30) | 11.2 (11.9) | 3.9 (2.4, 18.4) | Yes |
| Sit-to-Stand | 2 (80) | 66.8 (0) | 66.8 (66.8, 66.8) |  |
| Sling lift | 8 (20) | 66 (1.4) | 66.8 (65.6, 66.8) | Yes |
| Specialty walker<sup>b</sup> | 6 (40) | 80.4 (67.1) | 65.9 (46.4, 69.5) | Yes |
| Standard walker | 9 (10) | 55.1 (17.2) | 66.8 (47.5, 67.4) | Yes |
| Standing lift | 3 (70) | 66 (1.4) | 66.8 (65.6, 66.8) | Yes |
| Stretch band | 2 (80) | 40.6 (0.4) | 40.6 (40.5, 40.8) |  |
| Turning strap | 2 (80) | 50.3 (9.8) | 50.3 (46.8, 53.7) | Yes |

<sup>a</sup>Nonpharmacologic aids include ear plugs, eye covers, reading glasses, and/or amplifiers.
<sup>b</sup>walker that allows ambulation with fewer staff members; may include integrated features for oxygen tanks, portable ventilator/monitor, telescoping IV pole, seat flaps, leaning bar, among other features
*Abbreviations:* BMI = body mass index, m<sup>2</sup> = square meters, HOB = head of bed, IQR = interquartile range, PEEP = positive end expiratory pressure, SD = standard deviation

**Figure 1.**
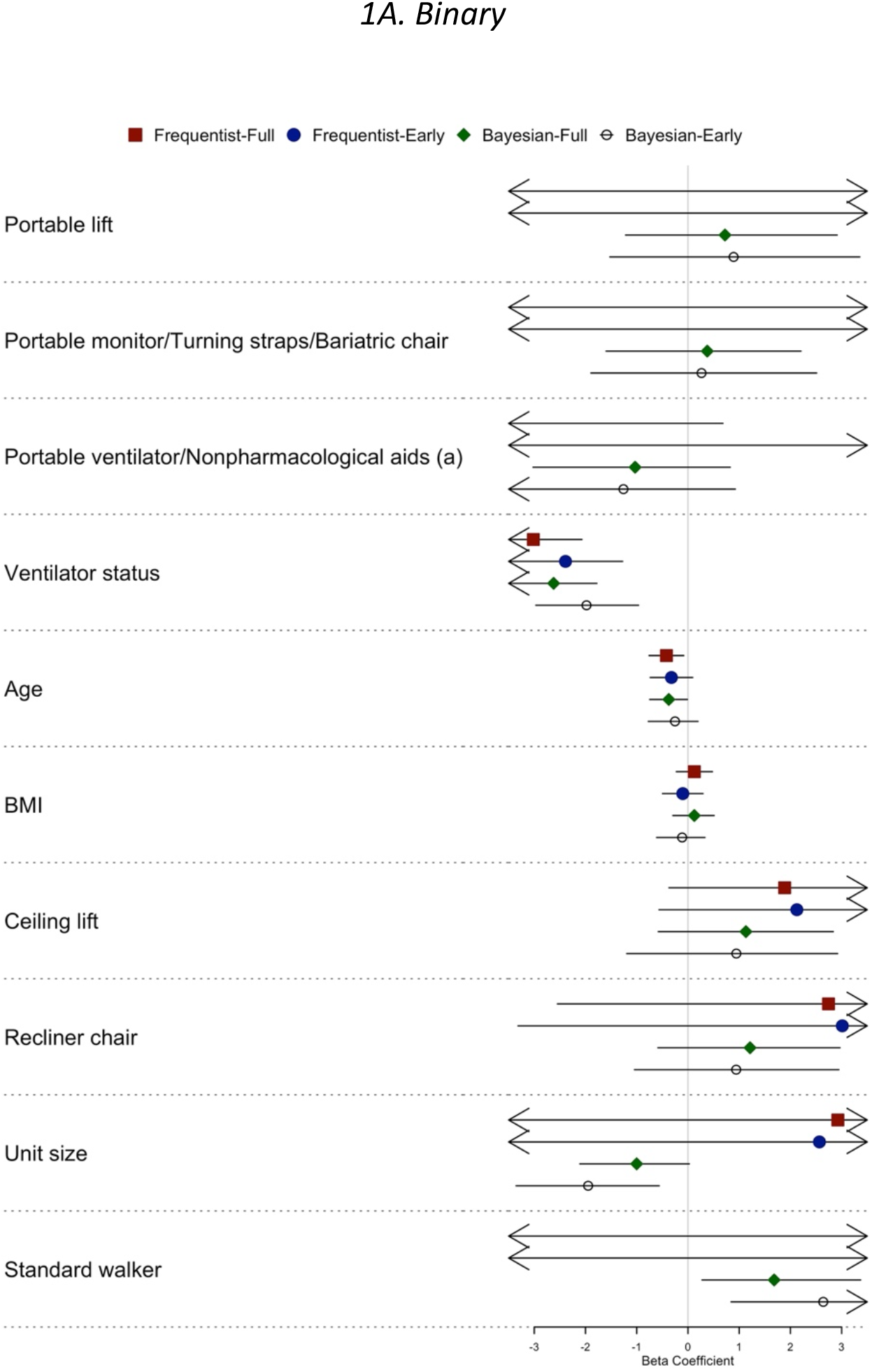
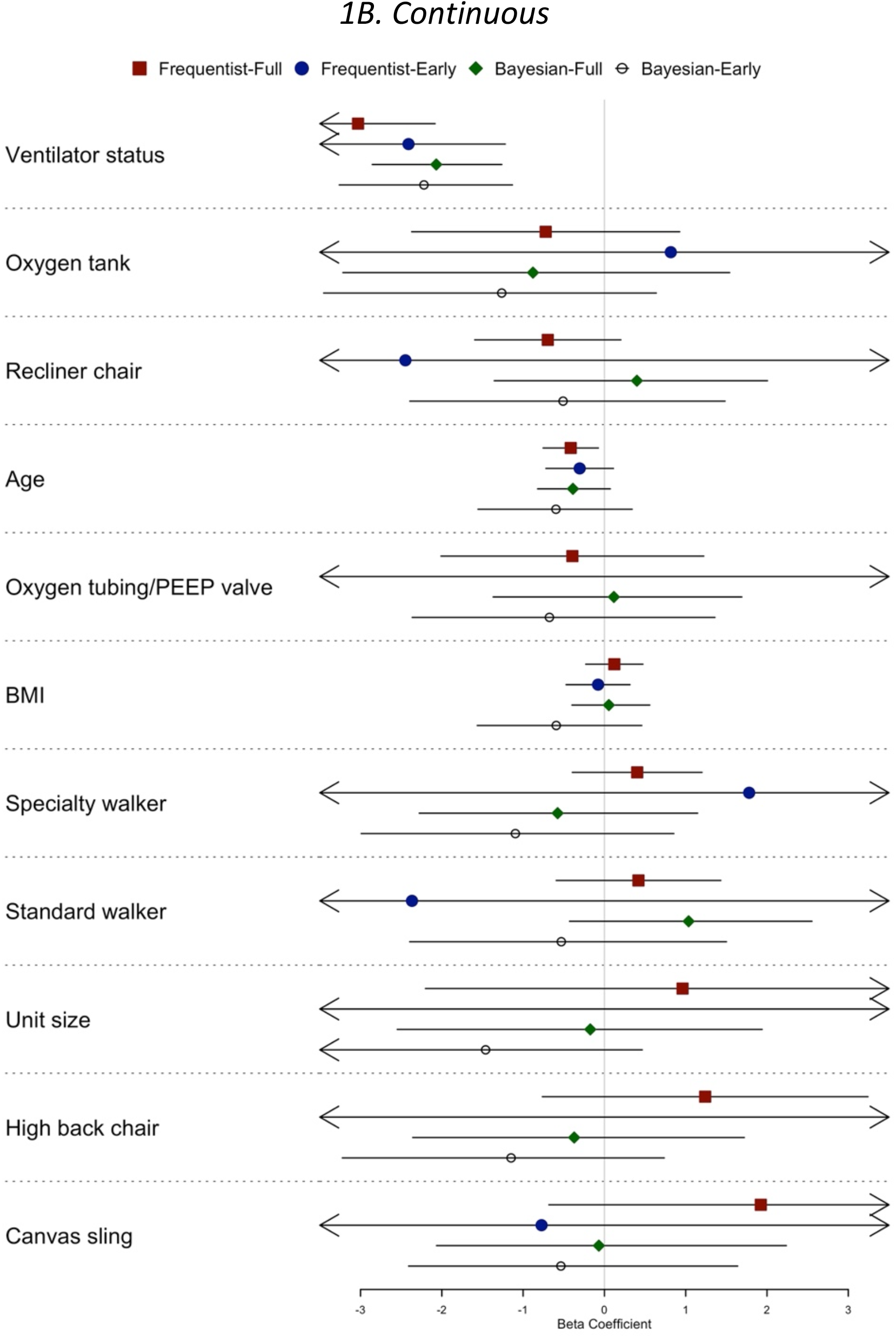
Beta coefficient means and 95% upper/lower limits for all 4 models using predictors for equipment, ranked in increasing order for mean of the frequentist approach for full adherence. (a) assumes binary imputation. (b) assumes continuous imputation.

For the Frequentist models, we used the lme4 package in R Studio (1.1.463, R Core Team, 2015) (https://cran.r-project.org/web/packages/lme4/vignettes/lmer.pdf). For the Bayesian models, we used PYMC3 (version 3.7) in Python (version 3.6.7). We used the No U-Turn Sampler suggested by Hoffman and Gelman (2014), which is a variation of the Hamiltonian Monte Carlo simulation method that allows for automatic tuning of step size and number.^24^ We developed models with uninformative priors (normal distributions with mean of 0 and standard deviations pulling from a half-Cauchy distribution with beta value of 5) and weakly informative priors (normal distributions with mean of -1 (for patient-level predictors and for equipment in the continuous models) or +1 (for equipment in the binary models) and standard deviations pulling from a half-Cauchy distribution with beta value of 2). We adjusted the number of tuning steps, iterations, and acceptance rates to optimize sampling chain convergence, Gelman-Rubin R-hat values, and Geweke z-scores.

## Results

The sample represents 751 patient observations across 105 patients within 10 different ICUs. The availability of ABCDEF-enhancing supplies and equipment ranged from 8-15 items across units. All units had electronic charts, bag valve masks, oxygen tubing, positive end expiratory pressure (PEEP) valves, and automated medication dispensing systems. The most commonly available bundle-enhancing items were standard walkers (n=9 units), ceiling lifts (n=7 units), and recliner chairs (n=7 units). The least commonly available (n=1-2 units) bundle-enhancing items were sit-to-stand aid, bariatric chair, portable ventilator, portable monitor, turning straps, stretch bands, and nonpharmacologic delirium aids. **Table 1** provides descriptive summaries for patient and unit level variables. Dot plot graphical comparisons of beta coefficients across the four best-fit models for each predictor type (i.e., binary and continuous) are found in **Figure 1a and 1b**, and corresponding numerical details are found in **Table 3** and **Table 4**. Beta coefficient estimate density plots for the best-fit Bayesian models are found in the Supplement (eFigures 1-4).

**Table 2.**
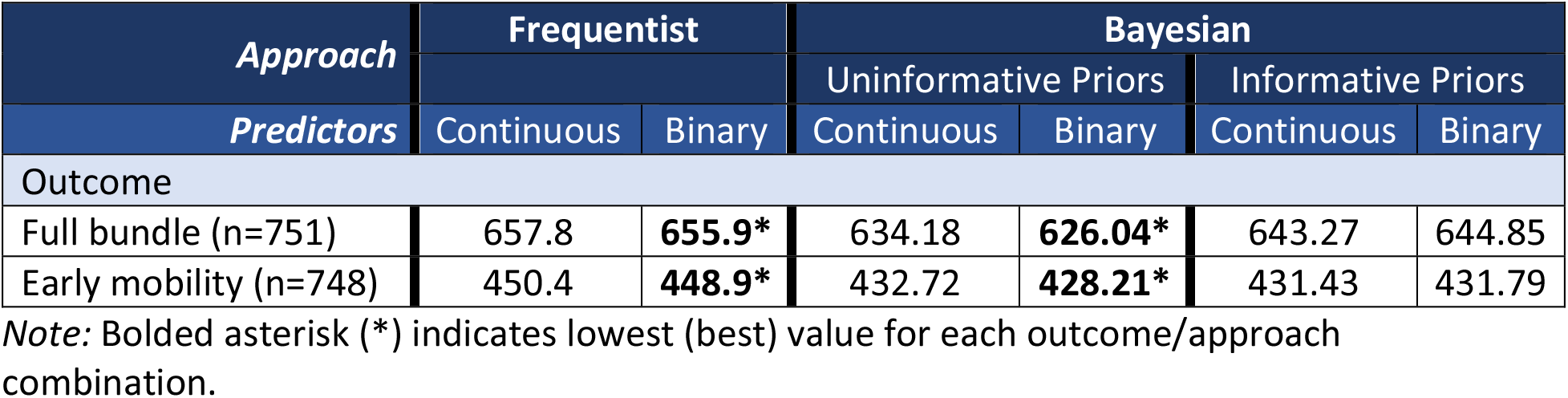
Comparison of best-fit models using AIC (Frequentist) or WAIC (Bayesian) values.

| *Approach* | Frequentist |  | Bayesian |  |  |  |
| --- | --- | --- | --- | --- | --- | --- |
|  |  |  | Uninformative Priors |  | Informative Priors |  |
| *Predictors* | Continuous | Binary | Continuous | Binary | Continuous | Binary |
| Outcome |  |  |  |  |  |  |
| Full bundle (n=751) | 657.8 | **655.9\*** | 634.18 | **626.04\*** | 643.27 | 644.85 |
| Early mobility (n=748) | 450.4 | **448.9\*** | 432.72 | **428.21\*** | 431.43 | 431.79 |

*Note:* Bolded asterisk (\*) indicates lowest (best) value for each outcome/approach combination.

**Table 3.** Beta coefficients for parameters of best-fit models examining adherence with full bundle.

| *Approach* | Frequentist |  | Bayesian |  |
| --- | --- | --- | --- | --- |
|  | Mean (standard deviation) |  | Mean (2.5% HPD, 97.5% HPD) |  |
| *Predictors* | Continuous | Binary | Continuous | Binary |
| **Daily Variable** |  |  |  |  |
| Ventilator status | -3.0 (0.5)*** | -3.0 (0.5)*** | -2.1 (-2.9,-1.3)* | -2.6 (-3.5,-1.8)* |
| **Patient Variables** |  |  |  |  |
| Age | -0.4 (0.2)* | -0.4 (0.2)* | -0.4 (-0.8,0.07) | -0.4 (-0.7,-0.01)* |
| BMI | 0.1 (0.2) | 0.1 (0.2) | 0.05 (-0.4,0.6) | 0.1 (-0.3,0.5) |
| **Unit Variables** |  |  |  |  |
| Unit size | 1.0 (1.6) | 2.9 (3.5) | -0.2 (-2.5,1.9) | -1.0 (-2.1,0.03) |
| Canvas sling | 1.9 (1.3) | n/a | -0.07 (-2.1,2.2) | n/a |
| Ceiling lift | n/a | 1.9 (1.2) | n/a | 1.1 (-0.6,2.8) |
| High back chair | 1.2 (1.0) | n/a | -0.4 (-2.4,1.7) | n/a |
| Oxygen tank | -0.7 (0.8) | n/a | -0.9 (-3.2,1.5) | n/a |
| Oxygen tubing/PEEP valve | -0.4 (0.8) | n/a | 0.1 (-1.4,1.7) | n/a |
| Portable lift | n/a | -12.0 (11.6) | n/a | 0.7 (-1.2,2.9) |
| Portable monitor/Turning straps/Bariatric chair | n/a | -8.7 (8.4) | n/a | 0.4 (-1.6,2.2) |
| Portable ventilator/Nonpharmacologic aids | n/a | -5.7 (3.3) | n/a | -1.0 (-3.0,0.8) |
| Recliner chair | -0.7 (0.5) | 2.7 (2.7) | 0.4 (-1.4,2.0) | 1.2 (-0.6,3.0) |
| Specialty walker | 0.4 (0.4) | n/a | -0.6 (-2.3,1.1) | n/a |
| Standard walker | 0.4 (0.5) | -5.3 (4.7) | 1.0 (-0.4,2.6) | 1.7 (0.3,3.4)* |

*Note:* \*\*\* p-value <0.001; \* p-value < 0.05 (Frequentist) or 95% HPD excludes 0 (Bayesian). Both Bayesian approaches assume uninformative priors.

**Table 4.** Beta coefficients for parameters of best-fit models examining adherence with early mobility component of bundle.

| *Approach* | **Frequentist** |  | **Bayesian** |  |
| --- | --- | --- | --- | --- |
|  | Mean (standard deviation) |  | Mean (2.5% HPD, 97.5% HPD) |  |
| *Predictors* | Continuous | Binary | Continuous | Binary |
| **Daily Variable** |  |  |  |  |
| Daily ventilator status | -2.4 (0.6)*** | -2.4 (0.6)*** | -2.2 (-3.3,-1.1)* | -2.0 (-3.0,-1.0)* |
| **Patient Variables** |  |  |  |  |
| Age | -0.3 (0.2) | -0.3 (0.2) | -0.6 (-1.6,0.3) | -0.3 (-0.8,0.2) |
| BMI | -0.1 (0.2) | -0.1 (0.2) | -0.6 (-1.6,0.5) | -0.1 (-0.6,0.3) |
| **Unit Variables** |  |  |  |  |
| Unit size | -9.5 (599.1) | 2.6 (4.1) | -1.5 (-3.7,0.5) | -1.9 (-3.4,-0.6)* |
| Canvas sling | -0.8 (133.7) | n/a | -0.5 (-2.4,1.6) | n/a |
| Ceiling lift | n/a | 2.1 (1.4) | n/a | 0.9 (-1.2,2.9) |
| High back chair | -4.9 (379.9) | n/a | -1.1 (-3.2,0.7) | n/a |
| Oxygen tank | 0.8 (71.9) | n/a | -1.3 (-3.5,0.6) | n/a |
| Oxygen tubing/PEEP valve | 4.1 (273.6) | n/a | -0.7 (-2.4,1.4) | n/a |
| Portable lift | n/a | -28.7 (65.4) | n/a | 0.9 (-1.5,3.4) |
| Portable monitor/Turning straps/Bariatric chair | n/a | -25.4 (65.3) | n/a | 0.3 (-1.9,2.5) |
| Portable ventilator | n/a | -21.9 (65.6) | n/a | -1.3 (-3.7,0.9) |
| Recliner chair | -2.4 (98.0) | 3.0 (3.2) | -0.5 (-2.4,1.5) | 0.9 (-1.0,2.9) |
| Specialty walker | 1.8 (88.4) | n/a | -1.1 (-3.0,0.9) | n/a |
| Standard walker | -2.4 (148.4) | 22.6 (65.4) | -0.5 (-2.4,1.5) | 2.6 (0.8,4.7)* |

Note: \*\*\* p-value <0.001; \* p-value < 0.05 (Frequentist) or 95% HPD excludes 0 (Bayesian).
Bayesian approaches assume uninformative priors for binary predictors and weakly-informative priors for continuous predictors.

### Full Bundle Adherence

In the Frequentist approach, both binary models for full bundle adherence and early mobility demonstrated better goodness of fit based on the Akaike information criterion (AIC) statistic (**Table 2**). Daily ventilator status and age were the only statistically significant predictors when modeling the outcome of full bundle adherence using various predictors in both the binary and continuous models (**Table 3**). Both variables had negative beta coefficients, indicating active mechanical ventilation and older age negatively influenced completion of the full bundle.

In the Bayesian approach, models with uninformative priors outperformed models with weakly-informative priors based on widely-applicable information criterion (WAIC) values (**Table 2**); therefore, primary findings are reported from models with uninformative priors. When using binary predictors, daily ventilator status, age, and availability of a standard walker had 95% high probability densities (HPDs) excluding 0 (**Table 3**). When using continuous predictors, daily ventilator status had 95% HPDs excluding 0. The beta coefficients for daily ventilator status and age were negative, indicating decreased full bundle adherence on days the patient is receiving mechanical ventilation and among older patients. The beta coefficient for walker availability was positive, indicating increased full bundle adherence on units with walkers.

### Early Mobility Adherence

For both binary and continuous models (**Table 4**), daily ventilator status was the only statistically significant predictor. Ventilator status had a negative regression weight, indicating active mechanical ventilation negatively influenced completion of early mobility.

In the Bayesian approach, models with uninformative priors outperformed models with weakly-informative priors when using binary predictors; conversely, models with weakly-informative priors outperformed models with uninformative priors when using continuous predictors (**Table 2**). When using binary predictors, daily ventilator status, a unit’s size, and availability of a walker had 95% HPDs excluding 0 (**Table 4**). When using continuous predictors, daily ventilator status had 95% HPDs excluding 0. Similar to the full bundle adherence models, the beta coefficients were negative for daily ventilator status and positive for standard walker availability. The beta coefficient for unit size was negative, indicating decreased early mobility adherence on larger units.

### Clinical Interpretation

Using variables that were statistically significant in the Frequentish approach or had HPDs excluding 0 in the Bayesian approach, one can convert the beta coefficients’ point estimates from all models into odds ratio ranges to aid interpretation, which we have done here. Being on the ventilator is independently associated with a patient being 8-20 times less likely to receive the full bundle and 7-11 times less likely to receive the early mobility component of the budle. Having a standard walker on the unit is associated with a patient being 5 times more likely to receive the fulle bundle and 13 times more likely to receive the early mobility component of the bundle. For every 1-year increase in a patient’s age the odds of receiving the full bundle are decreased by 33%. According to only 1 model, for every 1 meter^2^ increase in the unit size, the odds of receiving the early mobility component of the bundle are decreased by 85%.

## Discussion

We used multiple analytic models to examine the availability and accessibility of ABCDEF bundle-enhancing items in order to generate hypotheses about geospatial factors influencing bundle implementation. At the unit level, the presence of a standard walker was associated with full ABCDEF bundle and early mobility adherence. Larger unit size (m^2^) was associated with lower early mobility adherence. At the patient level, daily ventilator status was the only variable to consistently influence bundle adherence across all statistical models. Older age influenced the likelihood of receiving incomplete bundled care. Overall, the findings suggest the benefit of physical proximity to essential items for ABCDEF bundle and early mobility adherence.

Active mechanical ventilation was an important factor influencing both full bundle and early mobility adherence. Previous studies describe aversion to early mobility and ABCDEF bundle adherence due to perceived safety concerns or risk of adverse events (e.g., tube dislodgment, desaturation, fall). ^25, 26^ However, adverse events occurring with implementation of the ABCDEF bundle and related components are reportedly low.^5, 27, 28^ Similarly, increases in patient age were associated with decreases in full bundle adherence. In addition to safety concerns, this could be due to biased perceptions of older people as frail with diminished intrinsic capacity (i.e., the physical and mental capacity of an individual).^29^ Diverse trajectories of aging means there is no typical older person (e.g., 90-year-old marathon runner [high intrinsic capacity] vs. 60-year-old bedbound individual [frail]); thus, intrinsic capacity exists along a continuum.^29^ To address potential patient-level barriers to implementation, strategies may include interventions that overcome clinician hesitance with performing bundled care and implementing intrinsic capacity assessments at admission to develop care plans (e.g., mobility level goal) that maintain baseline function and avoid age-related bias.

At the unit level, our analysis builds on existing time-motion studies of the nurse work environment. In medical-surgical nursing units, when patient room assignments are clustered in close proximity and walking distances are minimized, the number of nurse-patient interactions increases.^30^ Similarly, our findings demonstrate that the geospatial location of mobility-related equipment could also be associated with use for full bundle adherence. Characteristics of the physical environment, both fixed (e.g., unit size) and modifiable (e.g., accessibility of a portable patient lift), may contribute to structure and process factors influencing full bundle implementation.

Strengths of our study comprise the heterogenous representation of medical and surgical ICUs from geographically diverse medical centers as well as the use of multiple analytic methods for triangulation. Slowly gaining popularity in the biomedical literature, Bayesian analyses have the advantage of incorporating prior knowledge and do not introduce error inflation during multiple testing. Further, our findings can be used as prior distributions in future studies leveraging Bayesian statistics, which effectively reduces the sample size needed in the future.

Our study has limitations that warrant consideration. The sample size of 10 ICUs from 6 academic medical centers does not represent the majority of ICU contexts, but does represent a mix of geographic regions in the Northeast, Southeast, Midwest, Southwest, and Pacific Northwest United States. The small sample size also limits our ability to make causal inference and resulted in sparse and collinear data for some pieces of equipment, necessitating their removal during final analyses. Evaluating bundle adherence by augmenting structural physical environment variables with process-related variables (e.g., team composition, staffing, and workflow) could also offer evidence-based insights for intervention development.^31^

## Conclusion

We identified unit- and patient-level barriers that may influence full bundle and early mobility implementation. There may be benefit to physical proximity of bundle-enhancing items, but we are limited in our ability to make causal inferences about the physical environment and adherence. There is benefit in testing implementation strategies that address hesitancy of performing bundled care with critically ill patients and applying early assessment of a patient’s intrinsic capacity. Future studies with larger sample sizes should explore how availability and accessibility (e.g., equipment location) could have implications to promote the implementation of evidence-based design.

## Data Availability

Data can be provided to interested readers upon request.

## Author Contributions

Drs. Werthman, Jeffery, Dietrich, and Boehm had full access to all of the data in the study and take responsibility for the integrity of the data and the accuracy of the data analysis.

*Concept and design:* Werthman, Jeffery, Mion, Boehm

*Acquisition, analysis, or interpretation of data:* Werthman, Jeffery, Boehm, Dietrich, Mion

*Drafting of the manuscript:* Werthman, Jeffery, Danesh, Boehm

*Critical revision of the manuscript for important intellectual content:* All

*Statistical analysis:* Werthman, Jeffery, Dietrich

*Obtained funding:* Boehm

*Administrative, technical, or material support:* Jeffery, Boehm

*Supervision:* Dietrich, Boehm

## Conflict of Interest Disclosures

LMB is currently receiving grant funding from the National Heart, Lung, and Blood Institute (#K12HL137943-01). ADJ received support for this work from the Agency for Healthcare Research and Quality (AHRQ) and the Patient-Centered Outcomes Research Institute (PCORI) under award number K12 HS026395. JW and ADJ received support from the resources and use of facilities at the Department of Veterans Affairs, Tennessee Valley Healthcare System. The content is solely the responsibility of the authors and does not necessarily represent the official views of AHRQ, PCORI, the Department of Veterans Affairs, or the United States Government. No other disclosures were reported.

## Funding/Support

This project was funded by an American Association of Critical-Care Nurses-Sigma Theta Tau International Critical Care Grant (#20170) and the Vanderbilt Institute for Clinical and Translational Research (UL1 TR000445 from National Center for Advancing Translational Sciences/National Institutes of Health).

## Role of the Funder/Sponsor

The funders had no role in the design and conduct of the study; collection, management, analysis, and interpretation of the data; preparation, review, or approval of the manuscript; and decision to submit the manuscript for publication.

## Additional Contributions

The authors would like to thank Dr. David Schlueter for contributing his knowledge and expertise during the technical design of the Bayesian modeling.

## Supplementary Material

## Supplementary Figures

**eFigure 1.**
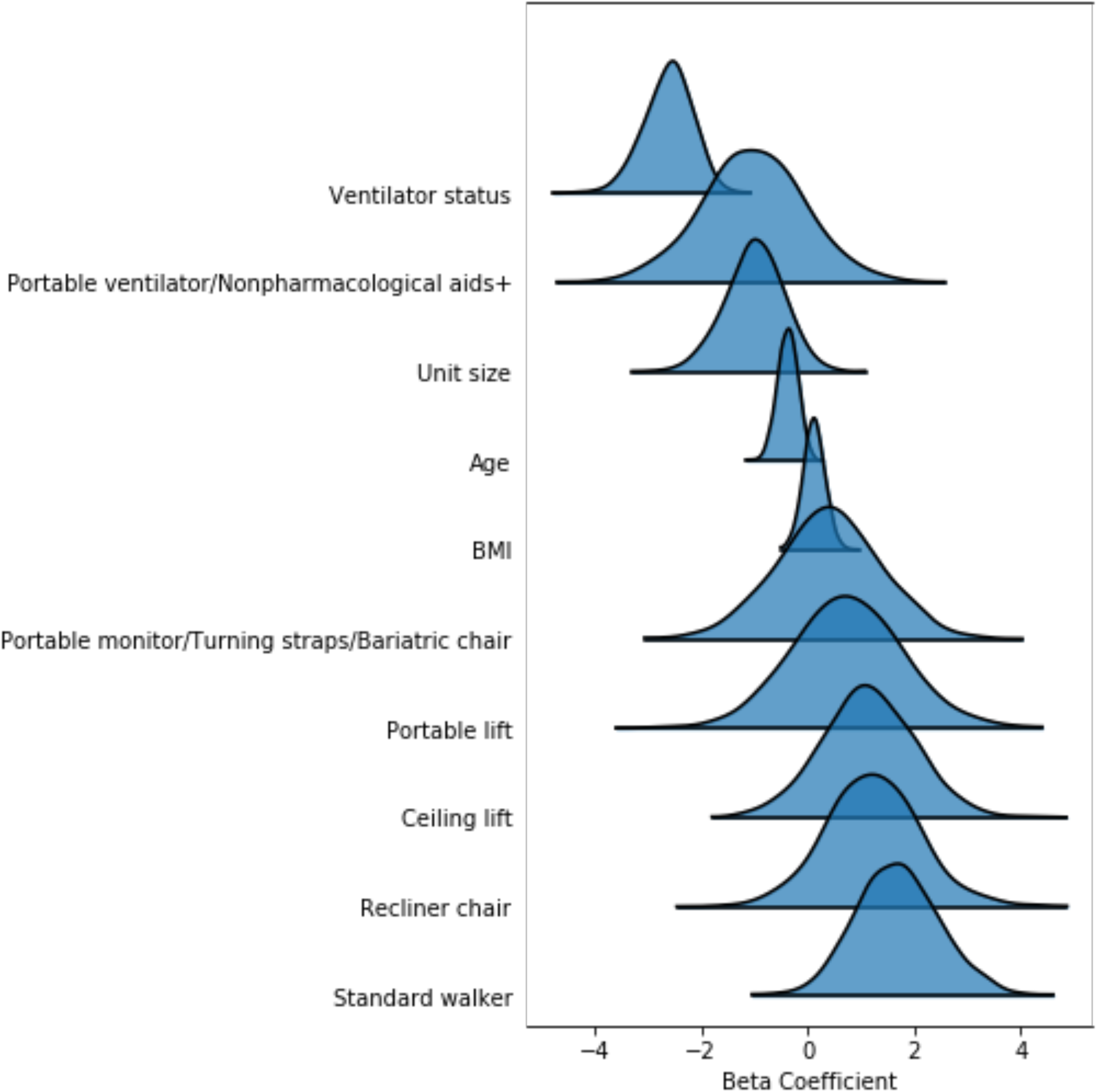
Highest density interval ridge plot representing beta coefficient estimates in the full bundle adherence Bayesian model with uninformative priors and binary predictors.

**eFigure 2.**
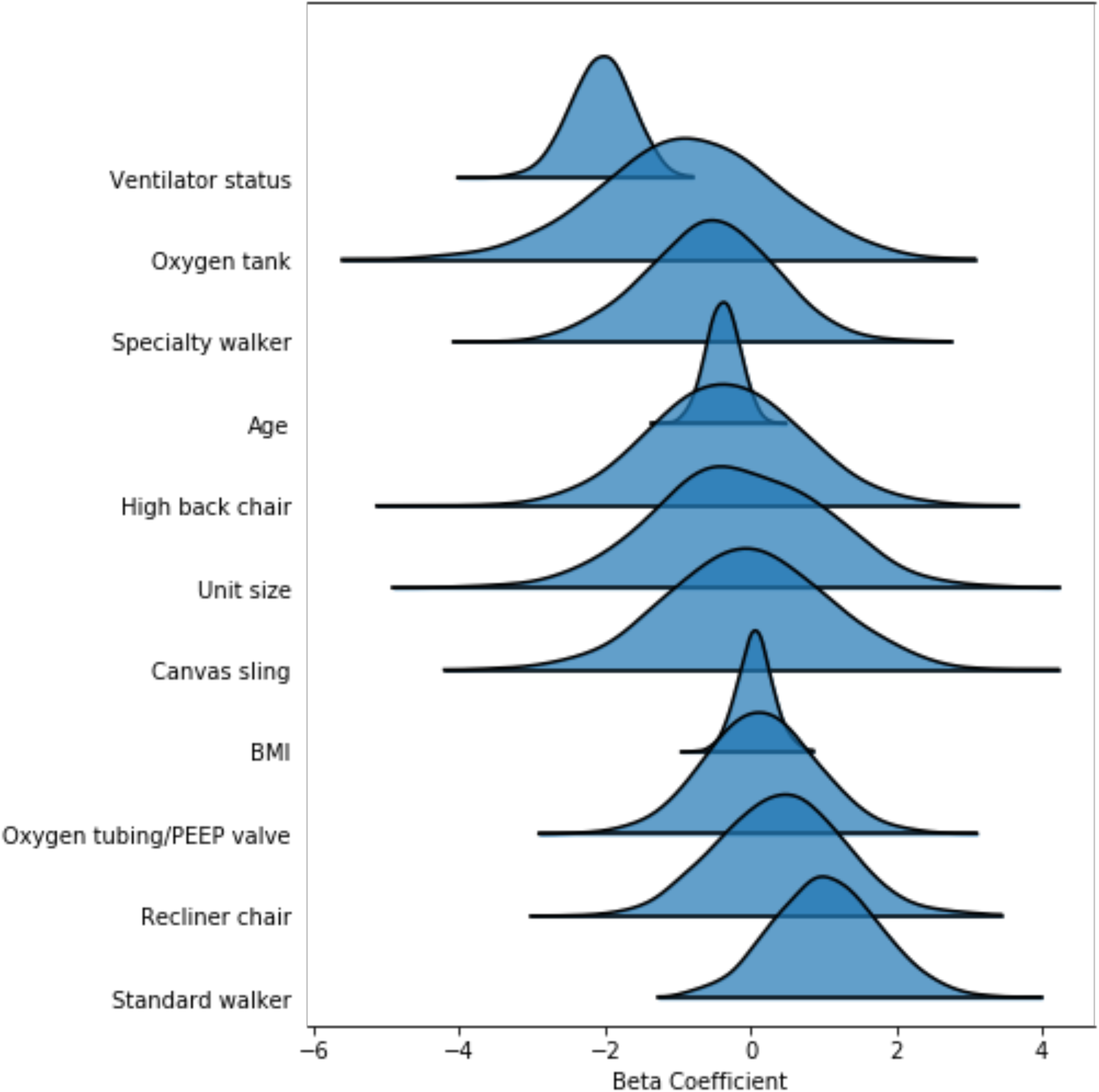
Highest density interval ridge plot representing beta coefficient estimates in the full bundle adherence Bayesian model with uninformative priors and continuous predictors.

**eFigure 3.**
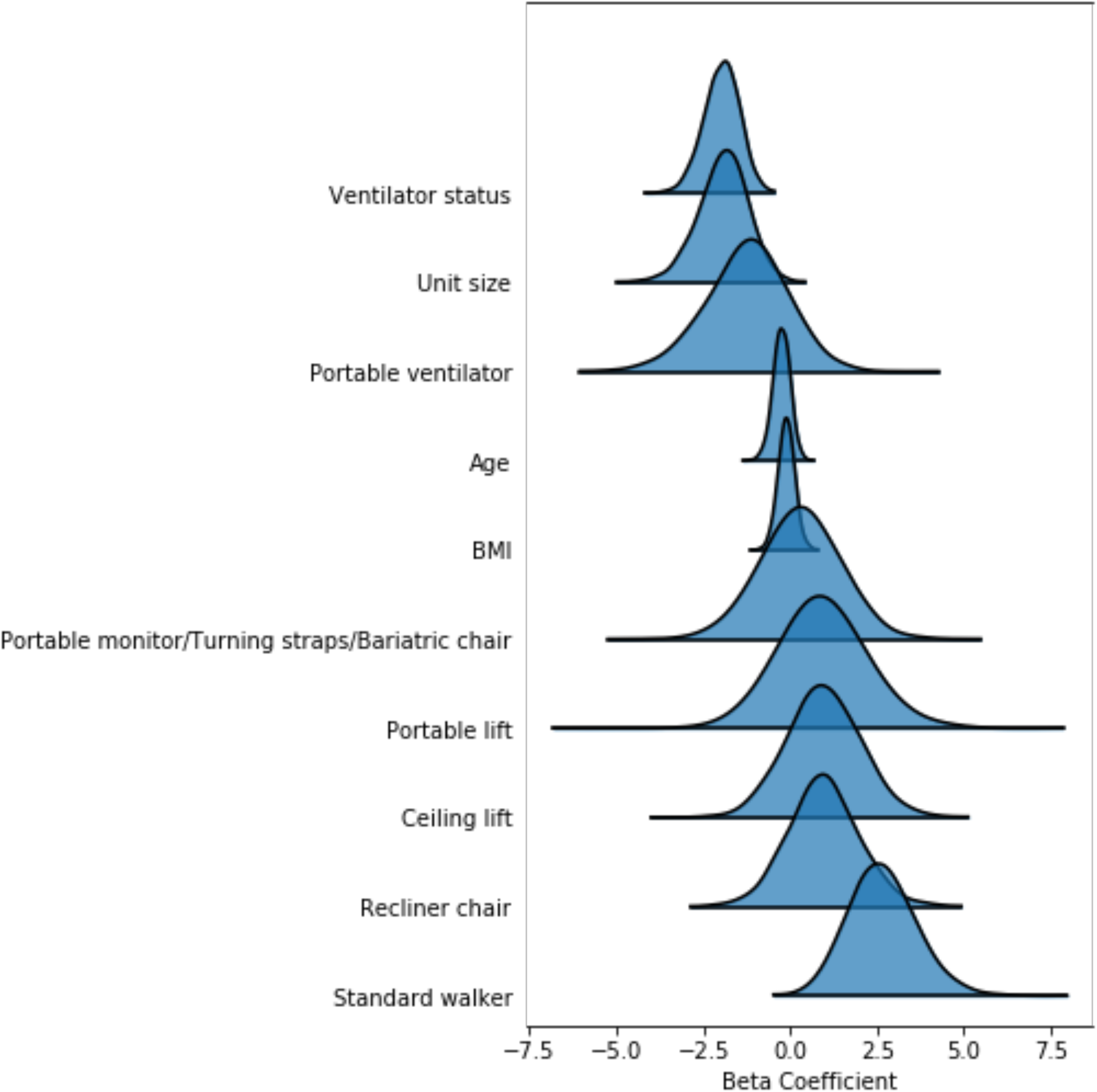
Highest density interval ridge plot representing beta coefficient estimates in the early mobility bundle adherence Bayesian model with uninformative priors and binary predictors.

**eFigure 4.**
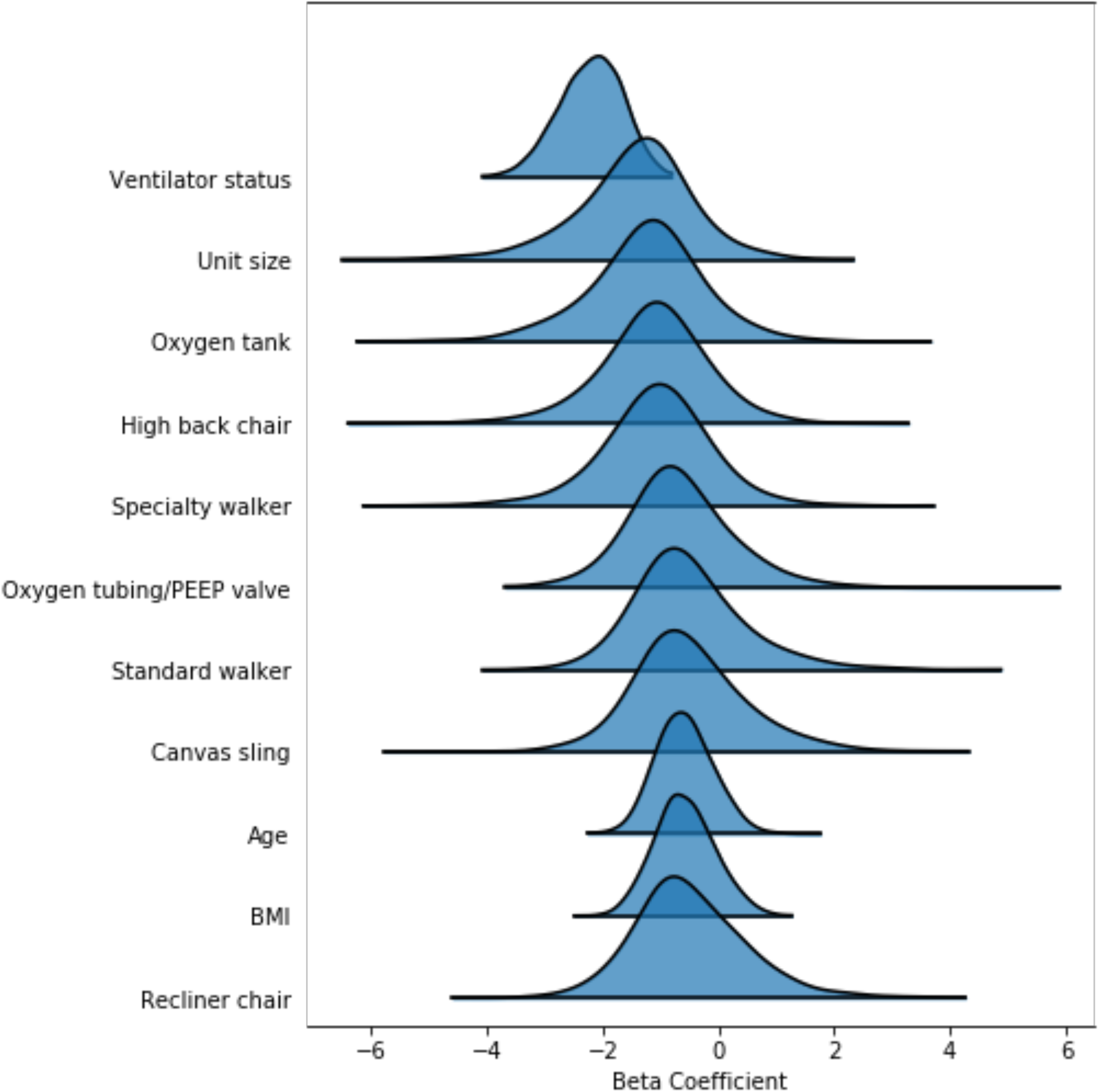
Highest density interval ridge plot representing beta coefficient estimates in the early mobility bundle adherence Bayesian model with weakly informative priors and continuous predictors.

**eTable 1.**
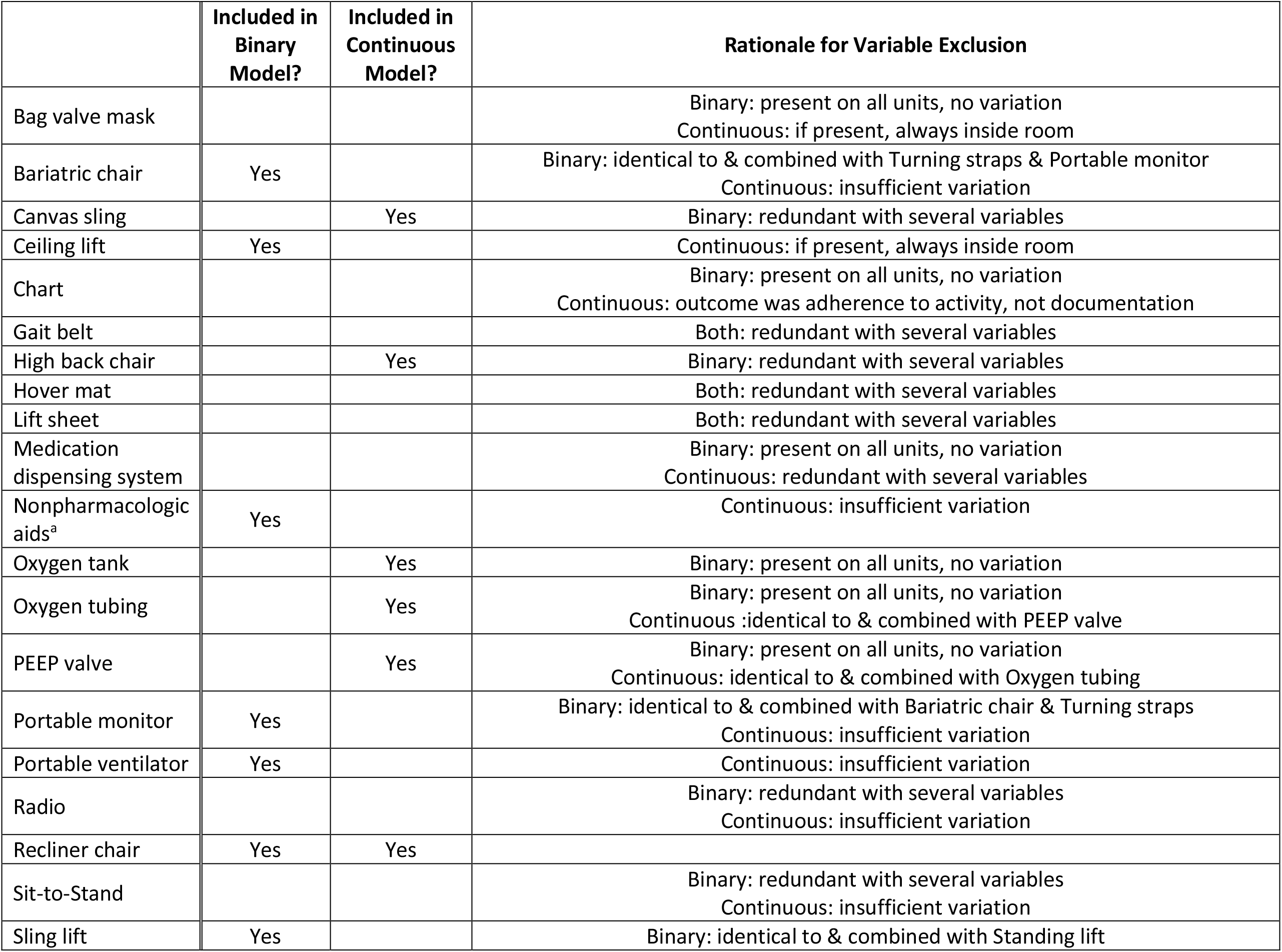
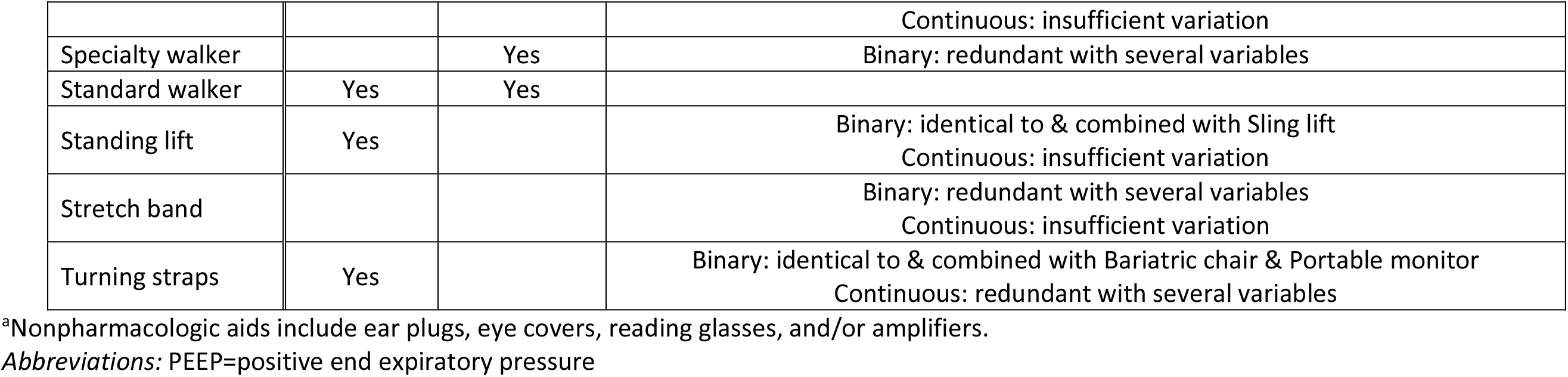
Equipment variable inclusion/exclusion following data-driven redundancy analysis with input from subject matter experts.

## Notes

### Competing Interest Statement

The authors have declared no competing interest.

### Author Declarations

This secondary, retrospective analysis had no specific institutional review board (IRB) oversight, as all data were already in aggregate form. For the primary data collection in the original studies, ethical approval was obtained from the IRB at each of the participating centers. The parent IRB approvals are held at Vanderbilt University under IRB# 040542 (MIND Study, NCT00096863) and IRB# 121380 (MENDS2 Study, NCT01739933). The following sub-sites also provided local IRB approval: University of California, San Francisco; Baton Rouge General Medical Center and Our Lady of The Lakes Regional Medical Center; Tufts Medical Center; Baystate Medical Center; Mission Hospital (Asheville, NC); Texas Health Harris Fort Worth; Baylor College of Medicine; Houston Methodist Hospital; University of Texas Health Science Center at San Antonio; University of Wisconsin; University of Iowa; University of North Carolina - Chapel Hill; Moses Cone (Greensboro, NC); & Saint Thomas Hospital (Nashville, TN).

